# Microbial-based yeast protein is similar to whey protein and greater than collagen hydrolysate in supporting whole-body protein synthesis as determined by the indicatory amino acid oxidation method: a randomized controlled trial

**DOI:** 10.64898/2026.06.13.26355601

**Authors:** Sabrina Tzivia Barsky, Hugo J. W. Fung, Nabil Bosco, Daniel R. Moore

**Affiliations:** Faculty of Kinesiology and Physical Education, University of Toronto, Toronto, ON, Canada; Lesaffre Institute of Science and Technology, Marcq-en-Barœul, France

**Keywords:** Protein quality, whole-body protein metabolism, amino acid oxidation, yeast-based protein, whey protein isolate, collagen protein

## Abstract

**Background:** Yeast (*Saccharomyces cerevisiae*) is a model organism in agricultural and industrial fermentation with nutritional benefits, yet less is understood about its nutritional value for supporting whole-body protein synthesis *in vivo*, and its comparison to animal-based protein.

**Objective:** This study aimed to determine the effect of microbial-based protein (yeast) compared to a high (whey) and low (collagen) quality animal-based protein on the ability to support whole-body protein synthesis using the indicator amino acid oxidation technique.

**Methods:** Thirteen healthy participants (M: *n*=6, 24±4 yr; F: *n*=7, 27±7 yr) consumed eight hourly yeast (Y), whey (W), or collagen (CH) protein beverages at 0.9 g·kg^−1^·d^−1^ supplemented with L-[1-^13^C]phenylalanine in a randomized, cross-over, counterbalanced design. Breath and urine were collected to measure fraction of expired ^13^CO_2_ (F^13^CO_2_) and [1-^13^C]Phe oxidation (PheOx) as inverse correlates of whole-body protein synthesis. An *a priori* 20% noninferiority margin was used to determine equivalency between protein sources. A Visual Analog Scale (VAS) was provided to assess fullness and hunger by protein sources.

**Results:** Protein source had no effect on F^13^CO_2_ (P=0.15) or PheRa (P=0.10). PheOx was greater in CH compared to both Y (14.94±2.16 vs. 12.93±2.80 μmol·kg BM^−1^·h^−1^, *P*<0.05) and W (14.94±2.16 vs. 12.50±2.75 μmol·kg BM^−1^·h^−1^, *P*<0.05) whereas there was no significant difference between Y and W (P=0.59). Change in PheOx (mean difference [90% CI]) between CH and W (2.44 μmol·kg BM^−1^·h^−1^ [0.98, 3.90], *P*<0.05) and between Y and W (0.44 μmol·kg BM^−1^·h^−1^ [−1.34, 2.21], P>0.99) confirmed respective inferiority and noninferiority to W compared to the 20% noninferiority margin (2.50 μmol·kg BM^−1^·h^−1^), while Y was superior to CH (−2.00 μmol·kg BM^−1^·h^−1^ [−3.01, −0.99], *P*<0.05). Protein source did not significantly influence satiety (P=0.32) or hunger scores (P=0.15).

**Conclusion:** EAA-enriched whey and yeast protein similarly reduced PheOx as compared to EAA-deficient collagen hydrolysate, suggesting these proteins were similarly superior in supporting whole-body protein synthesis.

## INTRODUCTION

Dietary protein is an essential macronutrient that provides the necessary substrates for physiological processes, including but not limited to growth and development of the musculoskeletal system, tissue and cellular maintenance, and gastrointestinal signaling of satiety (1). However, dietary proteins can differ in their digestibility as well as the content and ratio of essential amino acids (EAA), which can be captured in protein quality scoring metrics such as the digestible indispensable amino acid score (DIAAS) (2). In general, animal-based proteins are commonly enriched in EAA and have a high DIAAS (2). For example, whey protein is enriched in EAA and leucine (3), which contributes to its ability to robustly stimulate muscle and whole-body protein synthesis in humans (4). In contrast, plant-based proteins typically have a lower EAA content, may be limiting in one or more amino acids, and have poorer digestibility (3). However, alternative non-animal derived proteins that may be more sustainable or environmentally friendly are being explored for their ability to support human protein needs, such as those of microbial origin (5). It was recently demonstrated that yeast-based protein was of similar digestibility to whey and casein and has relatively high EAA content (∼47% compared to ∼49% in whey and casein) (6). Therefore, there is a need to evaluate the nutritional quality of novel microbial proteins such as yeast protein to support human amino acid requirements.

Non-invasive stable isotope tracer methods, such as the indicator amino acid oxidation (IAAO), have been useful in identifying the limiting amino acid which could limit protein synthesis responses (7,8). Specifically, the IAAO method uses the indicator amino acid, L-[1-^13^C]phenylalanine (1-[^13^C]Phe), to assess amino acid oxidation as a reciprocal for whole-body protein synthesis given the dual metabolic fates of EAA for protein synthesis or energy production (i.e. oxidation). As the IAAO determines the metabolic response to different levels or types of proteins, it represents a useful metabolic tool to evaluate the metabolic efficiency of dietary protein sources and their limiting amino acid(s) (9). Therefore, the overall objective of this investigation was to use the IAAO method to determine the ability of EAA-enriched (whey) or EAA-deficient (collagen) animal-derived proteins and a microbial (yeast) protein to attenuate whole-body phenylalanine oxidation as an inverse proxy for whole-body protein synthesis. We hypothesized that whey and yeast protein would support greater rates of whole-body protein synthesis (i.e. lower [1-^13^C]Phe oxidation) compared to collagen protein due to their relatively greater EAA content. Furthermore, we hypothesized that yeast protein would not be inferior to whey protein to support whole-body protein synthesis.

## SUBJECTS AND METHODS

### Participants and ethical statement

Thirteen healthy adults (pooled 6 M, 7 F) were informed of the study objectives, experimental procedures, and potential risks and gave written informed consent. Participants were required to complete the *Physical Activity Readiness Questionnaire* (PAR-Q+) and *International Physical Activity Questionnaire* (IPAQ 2002). To meet out inclusion criteria, eligible participants were required to be a healthy, recreationally active male or female between 18 and 45 years of age.

Females using monophasic combined oral contraceptives (COC) or those who reported a regular menstrual cycle (25-35 days) within the last 3 months were eligible for the study. Females recorded their menstrual cycle via the calendar method (10) to ensure all three metabolic trials were completed during the luteal phase. This allowed sex hormones (i.e. estrogen and progesterone) to remain consistent across trials (11), although fluctuations in sex hormones during different menstrual cycle phases do not seem to influence resting or exercise-induced muscle protein synthesis rates (12). Exclusion criteria included anabolic drug use, medications known to influence metabolism, or those who were on a vegan diet in the past 6 months. The studies were performed in accordance with the Declaration of Helsinki, and the study protocols were approved by the University of Toronto Delegated Research Ethics Board (REB#00048239). The study is registered at clinicaltrials.gov (NCT07148908).

**Table 1:** Subject characteristics of recreationally active, healthy men and women pooled.

|  | **Male** | **Female** | **Pooled** |
| --- | --- | --- | --- |
|  | *n*=6 | *n*=7 | *n*=13 |
| **Characteristics** | Mean ± SD | Mean ± SD | Mean ± SD |
| Age (years) | 24 ± 4 | 27 ± 7 | 26 ± 5 |
| Body Mass (kg) * | 82.5 ± 14.1 | 67.5 ± 13.8 | 73.9 ± 14.7 |
| Height (cm) | 184 ± 8 | 167 ± 10 | 175 ± 12 |
| BMI (kg/m<sup>2</sup>) | 24.2 ± 2.8 | 23.9 ± 3.1 | 23.8 ± 2.7 |
| FFM (kg) * | 68.8 ± 12.2 | 47.8 ± 7.9 | 57.6 ± 13.7 |
| Fat Mass (kg) * | 13.7 ± 6.0 | 19.8 ± 9.4 | 16.2 ± 7.7 |
| Body Fat % | 16.5 ± 6.3 | 28.2 ± 8.6 | 21.9 ± 8.8 |
| REE (kcal/day) ** | 2275 ± 545 | 1445 ± 207 | 1845 ± 540 |

\* Body mass, fat-free mass (FFM), and fat mass measured by BOD POD
\*\* Resting energy expenditure (REE) measured by indirect calorimetry

### Experimental Design

Following an overnight fast, participants reported to the Iovate/Muscletech Metabolism and Sports Science Lab at the University of Toronto to have their fat-free mass (FFM) and fat mass (FM) analyzed via air displacement plethysmography (BOD POD, Cosmed USA Inc., Chicago, IL). Participants’ resting energy expenditure (REE) was measured by indirect calorimetry using the TrueOne 2400 Metabolic Measurement System (Parvo, Utah, USA), and was used to produce their controlled study diet, which was consumed for 2 days prior to the metabolic trial day (1.5 × REE, 1.2 g protein·kg^−1^·d^−1^, 30% REE from dietary fats, and the remaining calories in the form of carbohydrates).

Following the 2-day controlled study diet, participants underwent an overnight fast (∼10 hr) and were instructed to avoid ingesting fluids prior to arrival on the study day. In a single-blind, randomized, counterbalanced design, participants consumed 1 of 3 dietary proteins sources (whey protein (W), yeast protein (Y), or collagen hydrolysate (CH)) in 8 hourly isocaloric and isonitrogenous meals, with each meal consisting of two-thirds of the participants total daily energy requirement (1.5x REE), measured by indirect calorimetry. The macronutrient breakdown of the metabolic trial day was 0.9 g protein·kg^−1^·d^−1^, 30% REE for dietary fats, and the remaining calories in the form of carbohydrates, consistent with previous IAAO studies at rest (13). The study day diet consisted of a protein- and AA-free powder (PFD1, Mead Johnson), maltodextrin (Maltodextrin, Canada Protein), grape-seed oil, one of three protein powders (whey protein [ALLMAX Nutrition, ON, Canada] and collagen hydrolysate [Great Lakes Wellness, Denver, CO, USA] were purchased commercially, while yeast protein was kindly provided by Lesaffre International) (**Table 2**), and protein-free cookies.

**Table 2:** Macronutrient and amino acid profile of animal- and microbial-based protein powders.

|  | CH |  | W |  | Y |  |
| --- | --- | --- | --- | --- | --- | --- |
| Calories (kcal) | 378 |  | 373 |  | 415 |  |
| Fat (g) | 0.38 |  | 0.51 |  | 8.79 |  |
| CHO (g) | <1.0 |  | 4.7 |  | 1.1 |  |
| Protein (g) | 93.6 |  | 87.4 |  | 82.9 |  |
| Amino Acid Composition |  |  |  |  |  |  |
| Essential | g/100g | % TAA | g/100g | % TAA | g/100g | % TAA |
| Isoleucine | 1.38 | 1.4 | 5.02 | 5.6 | 3.65 | 4.9 |
| Leucine | 2.67 | 2.6 | 8.58 | 9.5 | 6.12 | 8.2 |
| Valine | 2.29 | 2.3 | 4.59 | 5.1 | 4.2 | 5.6 |
| Histidine | 0.64 | 0.6 | 1.34 | 1.5 | 1.92 | 2.6 |
| Lysine | 3.45 | 3.4 | 8.38 | 9.3 | 6.79 | 9.1 |
| Methionine | 0.59 | 0.6 | 1.93 | 2.1 | 1.45 | 1.9 |
| Phenylalanine | 1.82 | 1.8 | 2.65 | 2.9 | 3.82 | 5.1 |
| Threonine | 1.79 | 1.8 | 6.43 | 7.1 | 4.08 | 5.5 |
| Tryptophan | <0.01 | 0.0 | 1.56 | 1.7 | 1.07 | 1.4 |
| Non-Essential |  |  |  |  |  |  |
| Aspartic Acid | 5.41 | 5.4 | 9.84 | 10.9 | 8.54 | 11.5 |
| Serine | 3.45 | 3.4 | 4.33 | 4.8 | 4.29 | 5.8 |
| Glutamic Acid | 9.93 | 9.9 | 15.4 | 17.1 | 8.71 | 11.7 |
| Glycine | 23.3 | 23.1 | 1.4 | 1.6 | 3.38 | 4.5 |
| Alanine | 8.91 | 8.8 | 4.65 | 5.2 | 4.56 | 6.1 |
| Tyrosine | 0.49 | 0.5 | 2.53 | 2.8 | 3.23 | 4.3 |
| Arginine | 7.6 | 7.5 | 1.64 | 1.8 | 4.33 | 5.8 |
| Proline | 14.1 | 14.0 | 7.36 | 8.2 | 3.66 | 4.9 |
| Hydroxyproline | 12.9 | 12.8 | <0.01 | 0.0 | <0.01 | 0.0 |
| Cysteine | 0.07 | 0.1 | 2.34 | 2.6 | 0.65 | 0.9 |
| Total AA | 100.79 | 100.0 | 89.97 | 100.0 | 74.45 | 100.0 |
| Total EAA | 14.63 | 14.5 | 40.48 | 45.0 | 33.10 | 44.5 |
| Total BCAA | 6.34 | 6.3 | 18.19 | 20.2 | 13.97 | 18.8 |

Protein analysis values are presented in grams per 100 grams protein powder with % of total amino acids (%TAA)
CH: collagen hydrolysate; W: whey protein; Y: yeast protein; AA: amino acids; EAA: essential amino acids; BCAA: branched chain amino acids (shaded)

Participants were given a Visual Analog Scale (VAS) to inquire about their appetite prior to the first drink, then again at 1-, 4-, and 8-hours following consumption of the first drink (14). The VAS was provided on a virtual survey portal, REDcap (Research Electronic Data Capture), hosted by the University of Toronto, and questions included: (1) how hungry do you feel?, (2) how full do you feel?, (3) how much of a desire do you have to eat?, and (4) how hungry do you anticipate being in the next 2 hours? VAS scores were collected digitally on a 1 to 100 linear scale. Respectively, hunger and satiety cues in the survey were measured as follows: Very hungry (1) to Not hungry at all (100); Not full (1) to Very full (100); Empty (1) to Satisfied (100); Cannot eat more (1) to Can eat more (100); and Very hungry in 2 hours (1) to Not hungry at all in 2 hours (100).

Provision of carbohydrate and fat content of the diets were adjusted (via maltodextrin and grape-seed oil, respectively) based on the macronutrient breakdown and nutritional profile of each commercially available protein (verified by Mérieux NutriSciences, Markham, ON, Canada) to keep the diets isoenergetic. Participants were not allowed to eat or drink anything else except for water *ad libitum*. The study days were separated by a minimum of 3 days to ensure tracer washout and to allow for at least one day of resumed habitual eating for improved control diet compliance. All subjects completed all 3 metabolic trials within 3 months.

### Tracer Protocol

An oral priming doses of 0.176 mg NaH^13^CO_2_·kg^−1^ body mass (99 atom% excess (APE); Cambridge Isotope Laboratories, Woburn, MA) and 0.66 mg L-[1-^13^C]phenylalanine (Phe) ·kg^−1^ body mass (99 atom% excess (APE); Cambridge Isotope laboratories, Woburn, MA) started with the fifth hourly meal. In addition, a dose of [1-^13^C]Phe (1.2 mg·kg^−1^·h^−1^) was started with the fifth meal and continued hourly for the remaining 4 h of the study. The quantity of [1-^13^C]Phe supplied during the last 4 h of the study was subtracted from the study diet meals to provide a total excess intake of 49 mg Phe·kg^−1^·d^−1^, with tyrosine also provided in excess within each hourly meal at 72 mg Tyr·kg^−1^·d^−1^ (13). This is to ensure metabolic partitioning of the phenylalanine carboxyl group to either synthesis or oxidation (15).

### Sample Collection

During each metabolic trial day, three baseline breath samples were collected at 45-, 30-, and 15-min before the priming dose of [1-^13^C]Phe was initiated at the fifth meal. Once isotopic steady state was attained 2 h following [1-^13^C]Phe tracer ingestion, a total of 6 breath samples were collected in duplicate, 45-, 30-, and 15-min before the seventh meal and similarly following the eighth meal. Breath samples were collected in 10 mL Vacutainers (BD, Franklin Lakes, NJ, USA) with a collection mechanism (Easy-Sampler, Quintron) that permitted the removal of dead-space air. Breath samples were stored at room temperature until analysis. Two baseline urine samples were collected at 20- and 40-min before the priming dose of [1-^13^C]Phe was initiated at the fifth meal, and three isotopic plateau urine samples were collected every 30-min following the seventh study meal. Urine samples were aliquoted for storage at −20°C before analysis. At each metabolic trial, the rate of rested carbon dioxide production (VCO_2_) was measured upon reaching metabolic and isotopic steady state after the sixth meal for 20-min via indirect calorimetry (TrueOne 2400 Metabolic Measurement System; Parvo, Utah, USA).

### Breath and Urine Sample Collection and Analysis

Breath samples were analyzed for expired ^13^CO_2_ enrichment by isotope ratio-mass spectrometry to determine [1-^13^C]Phe excretion with measured oxygen consumption (IDmicro Breath; Compact Science Systems, Staffordshire, UK) and a fed state bicarbonate retention factor of 0.82, as previously described (16,17). Briefly, breath samples were analyzed sequentially and with two technical replicates ordered in series. Atom % excess (APE) of ^13^CO_2_ was calculated from delta Pee Dee Belemite (PDB) values as follows:

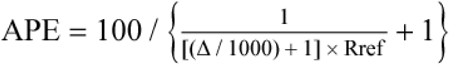

where Δ = delta PDB and Rref = 0.0111 for PDB (18).

Enrichments were expressed as APE compared with a reference standard of compressed CO_2_. Urinary [1-^13^C]Phe enrichment was measured by liquid chromatography tandem mass spectrometry (1290 HPLC, Agilent Technologies, Santa Clara, CA; Q-Trap MS, Sciex, Framingham, MA) by the Analytical Facility for Bioactive Molecules, The Hospital for Sick Children, Toronto, Canada. Isotopic enrichment was expressed as mole percent excess and calculated from peak area ratios at isotopic steady state both at baseline and plateau.

### Tracer kinetics

Phenylalanine flux or rate of appearance (PheRa, μmol·kg BM^−1^·h^−1^), ^13^CO_2_ excretion (F^13^CO_2_, μmol·kg BM^−1^·h^−1^), and phenylalanine oxidation (PheOx, μmol·kg BM^−1^·h^−1^) were calculated using the stochastic models described previously (13,19–21). Isotopic steady state in the tracer enrichment at baseline and plateau was represented as unchanging values of [^13^C]Phe in urine and ^13^CO_2_ in breath.

PheRa (μmol·kg BM^−1^·h^−1^) was calculated using the following equation:

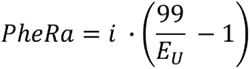

Where *i* represents the isotopic rate of [^13^C]Phe ingestion (μmol·kg BM^−1^·h^−1^) and E_u_ is the isotopic enrichments as a mole fraction in APE of urinary phenylalanine.

F^13^CO_2_ (μmol·kg BM^−1^·h^−1^) was calculated using the following equation:

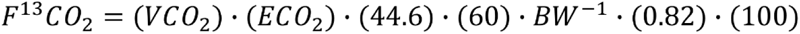

Where VCO_2_ is the rate of CO_2_ production (mL·min^−1^); ECO_2_ is the breath ^13^CO_2_ enrichment at isotopic steady state (APE); BW is body weight (kg). 44.6 μmol·kg^−1^·h^−1^ and 60 min·h^−1^ were constants used to convert F^13^CO_2_ to μmol·h^−1^. A factor of 0.82 was used to correct for CO_2_ retained in the bicarbonate pool in the fed state (19).

PheOx (μmol·kg BM^−1^·h^−1^) was calculated using the following equation:

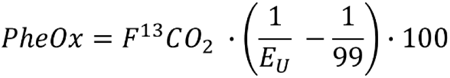

Where E_u_ is the isotopic enrichments as a mole fraction in APE of urinary phenylalanine and an estimate of intracellular enrichment (22).

### Statistics

Whey protein has a similar balanced and essential amino acid enriched profile as egg protein whereas collagen is deficient in most essential amino acids (**Table 2**) (3,23). Using the comparison of egg-modeled essential amino acids and non-essential amino acids (assumed similar comparison to whey and collagen), we estimated an expected effect size of 1.4 and standard deviation (SD) of 0.8 μmol·kg BM^−1^·h^−1^ for PheOx (24). With α=0.05 and β=0.9, n=8 participants are required to determine a difference between collagen and whey. To determine noninferiority of whey and yeast protein we used an *a priori* level of +20% PheOx, which based on our previous study approximated 1 μmol·kg BM^−1^·h^−1^. With α=0.05, β=0.9, and SD of 0.8 μmol·kg BM^−1^·h^−1^, n=13 was determined to be sufficient to accept or reject this noninferiority margin.

All analyses were completed using RStudio (2025.09.1, Build 401) with packages *ez*, *lme4*, *lmerTest*, *car,* and *eemeans*. Figures were produced by Prism9 (GraphPad Software Inc., San Diego, CA: Version 9.0.0). Data was analyzed using a within-subject approach with confirmation of normality by Shapiro-Wilk tests. Phenylalanine kinetics (i.e. F^13^CO_2_, PheRa, and PheOx) were analyzed by 1-way repeated measures ANOVA against protein source (i.e. whey, yeast, and collagen protein) with Greenhouse-Geisser epsilon correction applied to correct for sphericity. Appetite VAS scores were analyzed using a linear mixed-effects model (LMM) with timepoint (i.e. 0, 1, 3, and 7 hours), protein source, and their interaction included as fixed effects and participant ID included as a random intercept. VAS main and interactive effects were summarized by Type III ANOVA with Satterthwaite’s methods. *Post-hoc* pairwise comparisons were performed using estimated marginal means with Holm correction for multiple comparisons. The current study design was under-powered for a sex-based comparison, although an exploratory analysis using a LMM with protein source and sex as fixed effects revealed no differences by sex on any phenylalanine kinetics outcome, thus data analyses remain pooled by sex. Noninferiority of the test proteins (yeast and collagen) to the standard control (whey) was determined *a priori* by a margin of 20% of the primary outcome, PheOx. Noninferiority was analyzed by one-tail paired t-tests across a 90% confidence interval (CI) in sex-pooled data, with Bonferroni-correction applied for multiple comparisons. Statistical outliers were defined by a data point greater than the upper bound Q3 + 1.5 x IQR. Sensitivity analysis was performed with trials removed from any participants identified as outliers All data are presented as mean ± standard deviation (SD) with statistical significance set at α = 0.05.

## RESULTS

### Phenylalanine Kinetics

To minimize the effect of one extreme PheOx outlier (female participant during yeast trial day, Supplementary Data: **Figure S1C**), the data associated with this participant’s three metabolic trials was removed from the phenylalanine kinetics and inferiority data sets. A complete data set analysis (including the outlier participant) can be found in Supplementary Data, **Figure S1**.

PheOx was not different between conditions when all data were included (n=13; P = 0.07; **Figure S1C**). A sensitivity analysis was performed with this participant removed which revealed that PheOx was significantly different by protein source (*P*<0.05) (**Figure 1A**), with pairwise comparisons showing greater PheOx following CH consumption compared to that of W (14.94±2.16 vs. 12.50±2.75 μmol·kg BM^−1^·h^−1^, *P*<0.05) or Y (14.94±2.16 vs. 12.93±2.80μmol·kg BM^−1^·h^−1^, *P*<0.05). However, PheOx was not significantly different between W and Y consumption (12.50±2.75 vs. 12.93±2.8 μmol·kg BM^−1^·h^−1^, P=0.59). There were no significant effects of protein source on F^13^CO_2_ (P=0.15) or PheRa (P=0.10) (**Table 3**).

**Figure 1:**
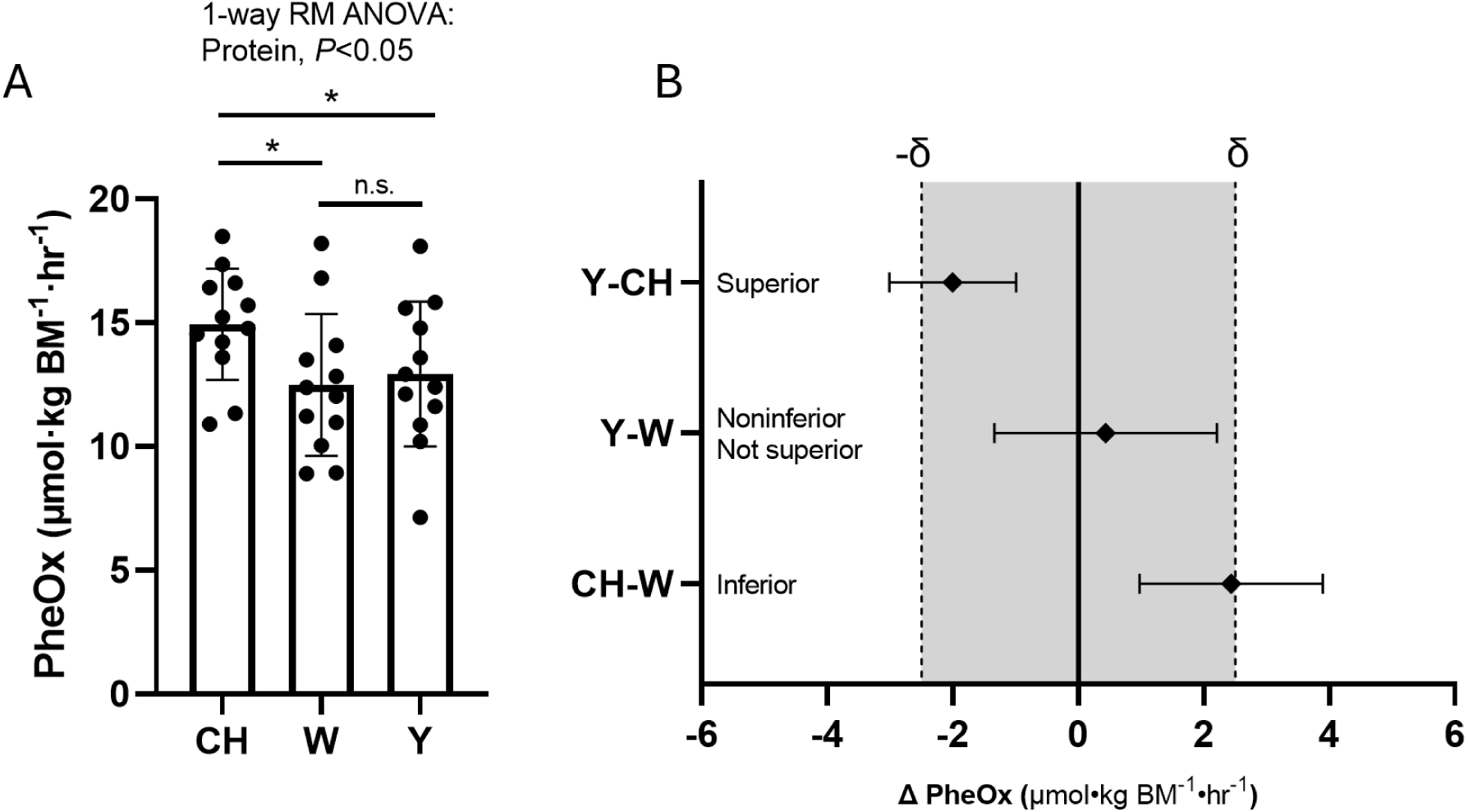
Yeast, but not collagen, is noninferior to whey in reducing PheOx. (**A**) Rate of phenylalanine oxidation (PheOx) (μmol·kg BM^−1^·hr^−1^) during the steady-state period following hourly collagen (CH), whey (W), or yeast (Y) ingestion at rest using a randomized crossover design. (**B**) Noninferiority of protein sources, collagen (CH) or yeast (Y) versus control comparator, whey (W) or CH, given change in PheOx. An outlier of PheOx in the yeast trial (22.99 μmol·kg BM^−1^·hr^−1^) was determined by any rate greater than Q3+1.5xIQR (21.55 μmol·kg BM^−1^·hr^−1^) and all subsequent trials of this outlier were removed. Data are presented as means ± SD (n = 12 per group). (**A**) Data are analyzed by a 1-way repeated-measures ANOVA with a pairwise *post-hoc* estimated marginal means test with Holm correction for multiple comparisons. Significance denoted by *, *P*<0.05. (**B**) Data analyzed by Bonferroni-corrected one-tailed paired t-tests, with W set as control comparator. Noninferiority against whey is accepted when the upper bound of the 90% CI in change of PheOx is lower than the predetermined 20% noninferiority margin, with superiority accepted when lower bound crosses the superiority margin (+/-δ, +/-2.50 μmol·kg BM^−1^·hr^−1^, *grey shading*).

**Table 3:** Phenylalanine flux between protein sources (collagen (CH), whey (W), or yeast (Y)). Fraction of expired ^13^CO_2_ (μmol·kg BM^−1^·hr^−1^), and rate of phenylalanine appearance (PheRa) (μmol·kg BM^−1^·hr^−1^) are presented as means ± SD (n = 12 per group, all outlier trials removed). Data are analyzed by a 1-way repeated-measures ANOVA. There are no main effects of protein source on F^13^CO_2_ (P=0.15) or PheRa (P=0.10).

|  | CH | W | Y | F-statistic | P-value |
| --- | --- | --- | --- | --- | --- |
| | Mean ± SD | Mean ± SD | Mean ± SD | ($\epsilon[k-1]$, $\epsilon[[k-1][n-1]]$) | |
| $\text{F}^{13}\text{CO}_2$<br>$\mu\text{mol}\cdot\text{BM}^{-1}\cdot\text{hr}^{-1}$ | $1.96 \pm 0.39$ | $1.74 \pm 0.27$ | $1.83 \pm 0.39$ | $F(1.81, 19.93) = 2.110$ | 0.15 |
| PheRa<br>$\mu\text{mol}\cdot\text{BM}^{-1}\cdot\text{hr}^{-1}$ | $55.31 \pm 7.48$ | $51.43 \pm 8.41$ | $50.74 \pm 5.80$ | $F(1.78, 19.52) = 2.670$ | 0.10 |

$k$ , Total treatment groups
$n$ , Total sample size
$\epsilon$ , Greenhouse-Geisser Epsilon correction

### Yeast is noninferior to whey but superior to collagen

To understand inferiority or superiority of Y to CH, and Y or CH in comparison to the standard W protein, on the primary outcome, PheOx, there was a predetermined margin of inferiority set at 20%. Given that PheOx is inversely proportional to whole-body protein synthesis, a lower PheOx describes greater incorporation of the protein source into tissue pools, rather than use for energy substrate metabolism via oxidation. In sex pooled data (**Figure 1B**), change in PheOx (mean difference [90% CI]) was significant between CH and W (2.44 μmol·kg BM^−1^·h^−1^ [0.98, 3.90], *P*<0.05), but insignificant between Y and W (0.44 μmol·kg BM^−1^·h^−1^ [−1.34, 2.21],

P>0.99). Neither test protein versus whey CI upper bound crosses the 0 change across the x-axis, indicating that neither test protein is superior to whey on eliciting a lower PheOx, and thus protein synthesis response. However, Y was found to be superior to CH in lowering PheOx rates (−2.00 μmol·kg BM^−1^·h^−1^ [−3.01, −0.99], *P*<0.05), with the 90% CI spread crossing the negative margin of inferiority (i.e. differences between PheOx are larger between the protein sources).

Noninferiority of Y to W is observed as the entire CI spread lies behind the margin of inferiority (i.e. differences between PheOx are minimal between the protein sources), with CH showing inferiority against W, as the upper bound of the 90% CI crosses the margin of inferiority (i.e. differences between PheOx are larger between the protein sources). Inclusion of the statistical outlier resulted in CH remaining inferior to W whereas the noninferiority of Y to W was inconclusive and Y did not retain superiority to CH (**Figure S1D**).

### Appetite scores were unchanged by protein source

Given the higher fat content in a serving of yeast protein compared to whey and collagen (**Table 2**), we predicted that appetite and satiety would be reduced and increased respectively across the trial day in a protein-dependent manner. However, there were no statistical main effects of protein source on any current hunger (P=0.15), fullness (P=0.18), satiety (P=0.32), desire to eat (P=0.60), or prospective hunger (P=0.27) scores (**Figure 2A-E**). Time presented a significant effect on fullness (**Figure 2B**, *P*<0.001) and satiety (**Figure 2C**, *P*<0.001) such that feeding throughout the trial day increased both reported metrics of fullness with no difference between conditions (*P*≥0.18). *Post-hoc* comparisons revealed significant increases in fullness between timepoints 0- and 3-hours (35 ± 20 vs. 47 ± 17; *P*<0.01), 1- and 7-hours (41 ± 17 vs. 51 ± 20; *P*<0.05), and 0- and 7-hours (35 ± 20 vs. 51 ± 20; *P*<0.001). Similarly, pairwise comparisons by time showed increased subjective satiety between timepoints 0- and 3-hours (35 ± 19 vs. 45 ± 14; *P*<0.05), 1- and 7-hours (39 ± 14 vs. 48 ± 19; *P*<0.05), and 0- and 7-hours (35 ± 19 vs. 48 ± 19; *P*<0.01). Prospective food consumption score (i.e. higher score indicates satiety) presented a main effect of time (**Figure 2E**, *P*<0.001), with *post-hoc* comparisons showing less prospective hunger throughout the trial day, as revealed between 0- and 1-hour (20 ± 12 vs. 30 ± 16; *P*<0.05), 0- and 3-hours (20 ± 12 vs. 36 ± 22; *P*<0.001), and 0- and 7-hours (20 ± 12 vs. 32 ± 23; *P*<0.001). Altogether, VAS data suggests that greater fullness and less prospective hunger were associated with drink consumption throughout the trial in a protein-independent manner.

**Figure 2:**
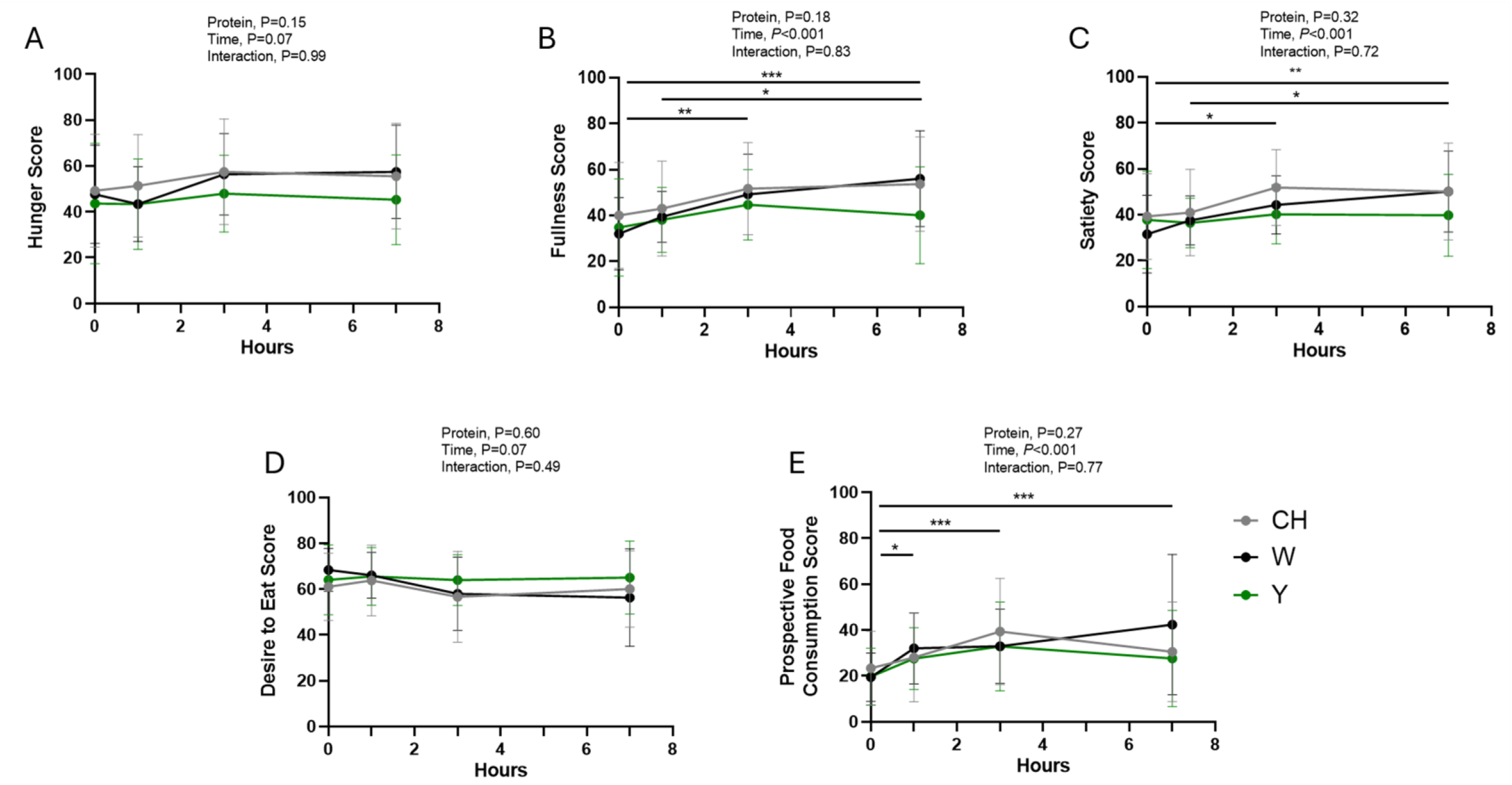
Protein source does not affect appetite or satiety over trial day. Appetite and satiety scores as measured by Visual Analog Scale (VAS) at 0-, 1-, 3-, and 7-hours following consumption of the first (CH, *grey*), whey (W, *black*), or yeast (Y, *green*) protein beverage at rest in a randomized crossover design. (**A**) Hunger Score, (**B**) Fullness Score, (**C**) Satiety Score, (**D**) Desire to Eat Score, and (**E**) Prospective Consumption Score were measured prior to the consumption of the drink on a sliding scale from 1-100. Data are presented as mean ± SD (n = 13 per group). Data are analyzed by linear mixed effects model and followed by *post-hoc* pairwise estimated marginal means test with Holm correction for multiple comparisons. There are no main effects of protein on any appetite or satiety score, however time yields a main effect on fullness (*P*<0.001), satiety (*P*<0.001), and prospective hunger scores (*P*<0.001), with significant *post-hoc* comparisons between timepoints denoted by *, *P*<0.05; **,*P*<0.01; ***, *P*<0.001.

## DISCUSSION

Using the IAAO technique, we demonstrated that yeast and whey protein isolates induced a lower phenylalanine oxidation than an isocaloric and isonitrogenous hydrolyzed collagen protein when consumed at an intake approximating the IAAO-derived EAR for protein.

Furthermore, according to an *a priori* 20% bio-equivalency threshold, yeast protein is not inferior to whey protein in attenuating phenylalanine oxidation and is superior to collagen. Collectively, our results suggest that yeast and whey protein similarly support whole-body protein synthesis in healthy adults.

We have previously used the IAAO to determine the most limiting amino acids in the diets of endurance athletes by providing a sub-optimal intake and adding back in various combinations of EAA and NEAA (24,25). We adapted this approach for the present study by providing a single isolated protein as the sole nitrogen source at an intake that would approximate the EAR in a healthy young population (13). Providing an intake around the EAR would allow for relative differences in protein quality to be elucidated by providing insight into the metabolic availability of the test proteins to support whole-body protein synthesis (26). Consistent with its low DIAAS score and primarily NEAA profile (2), collagen protein ingestion was associated with greater phenylalanine oxidation than whey, suggesting a limitation in one or many amino acids to support whole-body protein synthesis. While we cannot determine what amino acid(s) limited the anabolic potential of collagen, independent analysis of our proteins revealed a pronounced deficiency of EAA in collagen relative to whey (∼14.5 vs ∼45%, respectively) and presume these anabolic amino acids are primarily responsible (27).

Importantly, we purposely utilized a collagen hydrolysate to minimize potential differences in digestibility between whey and native collagen (28). The novel finding for the present study, however, was the lower phenylalanine oxidation in yeast compared to collagen, which would be consistent with its relatively high (∼44%) EAA content to support whole-body protein synthesis. Therefore, our results are consistent with EAA-enriched proteins such as whey and yeast as being superior to EAA-deficient proteins such as collagen in providing the requisite amino acids to support body protein synthesis.

Our secondary objective to determine how yeast protein compared to whey protein in supporting whole-body protein synthesis revealed that it was within our *a priori* identified noninferiority margin of 20%. Yeast and whey protein have previously been shown to have similar digestibility by multiple *in vitro* models with their DIAAS greater than 0.97 (6,29).

Furthermore, when provided at the EAR for protein according to the IAAO as in the present study, both yeast and whey provide greater than the average daily requirement of leucine (30), lysine (31), threonine (32), tryptophan (33), and the sulfur amino acids (34), all of which collagen is incidentally deficient in. Therefore, the noninferiority of yeast protein highlights that it has a similarly high metabolic availability (presumably of EAA) as whey protein.

Food ingestion, timing of consumption bouts, bolus size, and macronutrient content are factors which regulate acute satiety, marked by the release of gastrointestinal incretins, notably glucagon-like peptide 1 (GLP-1) (35). In an *in vitro* model of digestion, yeast protein resulted in greater GLP-1 secretion in STC-1 enteroendocrine cells compared to casein, suggesting that it may have greater impact on satiety *in vivo* (6). As a tertiary outcome, we also assessed the impact of protein source on subjective sensations of appetite and fullness based on our prediction that the higher fat profile of yeast (∼9g per 100g serving) compared to both whey (∼0.5g per 100g serving) and collagen (∼0.4g per 100g serving) would contribute to increased subjective satiety. Our findings suggest that neither protein source, which were provided in an isocaloric and isonitrogenous manner, impacted hunger and satiety ratings, and instead, satiety was mostly affected by consistent eating throughout the trial day. This would be consistent with GLP-1 release and its subsequent response on perceived satiety having a stronger association with meal size than macronutrient composition (36), which was not controlled in the current study. For example, healthy adults provided isocaloric and isovolemic meals, postprandial GLP-1 secretion shows mixed magnitude of response based on macronutrient profiles, with greater increases following higher protein content compared to higher fats or carbohydrates (37) or equivalent increases in both high protein and high fat drinks (38). Overall, our use of a modified IAAO technique resulted in several required methodologies that would likely affect satiety to a stronger degree than the macronutrient profiles of isonitrogenous and isocaloric meals, including (i) hourly liquid bolus feedings to maintain phenylalanine pool steady-state assumptions, (ii) total energy provided in excess to avoid inducing any degree of catabolism (i.e. 1.5xREE with 30% fat contribution), and (iii) water provided *ad libitum* throughout trial days.

Traditional IAAO methodology using crystalline amino acids as the ‘protein’ source often relies on phenylalanine excretion (F^13^CO_2_) to estimate the difference between levels of protein to support whole-body protein synthesis (i.e. the reciprocal of the oxidation of the indicator amino acid). However, this approach does not consider whether the pool size of phenylalanine (as reflected by changes in urinary enrichment and phenylalanine flux) is influenced by the test protein intake. While we did not observe any statistical differences in phenylalanine flux between proteins, there were visual trends that, despite providing equivalent total phenylalanine intake in excess of the daily requirement, the test proteins may have elicited subtle differences in intracellular phenylalanine enrichment that should be accounted for.

Therefore, we took the approach to determine the anabolic potential of the test proteins based on differences in phenylalanine oxidation, which incorporates both breath ^13^CO_2_ enrichment (i.e. F^13^CO_2_) and urinary phenylalanine enrichment (i.e. phenylalanine flux) into this physiological outcome of tracer oxidation and was the primary outcome to which our sample size calculation was based on.

In summary, we demonstrate that the EAA-enriched proteins whey and yeast result in a lower oxidation of our indicator amino acid than the EAA-deficient hydrolyzed collagen protein. Moreover, yeast and whey protein were similar in their ability to reduce phenylalanine oxidation, which highlights these proteins as being of similarly high quality to support whole-body protein synthesis. However, when consumed with sufficient energy and in small hourly meals (consistent with IAAO methodology), protein source did not influence subjective markers of satiety.

## DATA AVAILABILITY

Study data is available by reasonable request to the corresponding author.

## ETHICS STATEMENT

All participants were informed of the purpose of the study, experimental procedures, and potential risks and gave written informed consent. The studies were performed in accordance with the Declaration of Helsinki, and the study protocols were approved by the University of Toronto Delegated Research Ethics Board (REB#00048239). The study is registered at clinicaltrials.gov (NCT07148908).

## AUTHOR CONTRIBUTIONS

SB and DM conceptualized the research. SB conducted the research and collected all data. SB and HF analyzed the data. SB drafted the manuscript with input from HF and NB, and DM provided critical revisions to the manuscript. All authors have read and approved the final manuscript.

## FUNDING

Funding provided by Lesaffre International and administered by University of Toronto to DM.

## CONFLICT OF INTEREST

The authors declare that NB is an employee of Lesaffre International which may be considered a potential source of competing interest.

## SUPPLEMENTARY FIGURES

**Figure S1:**
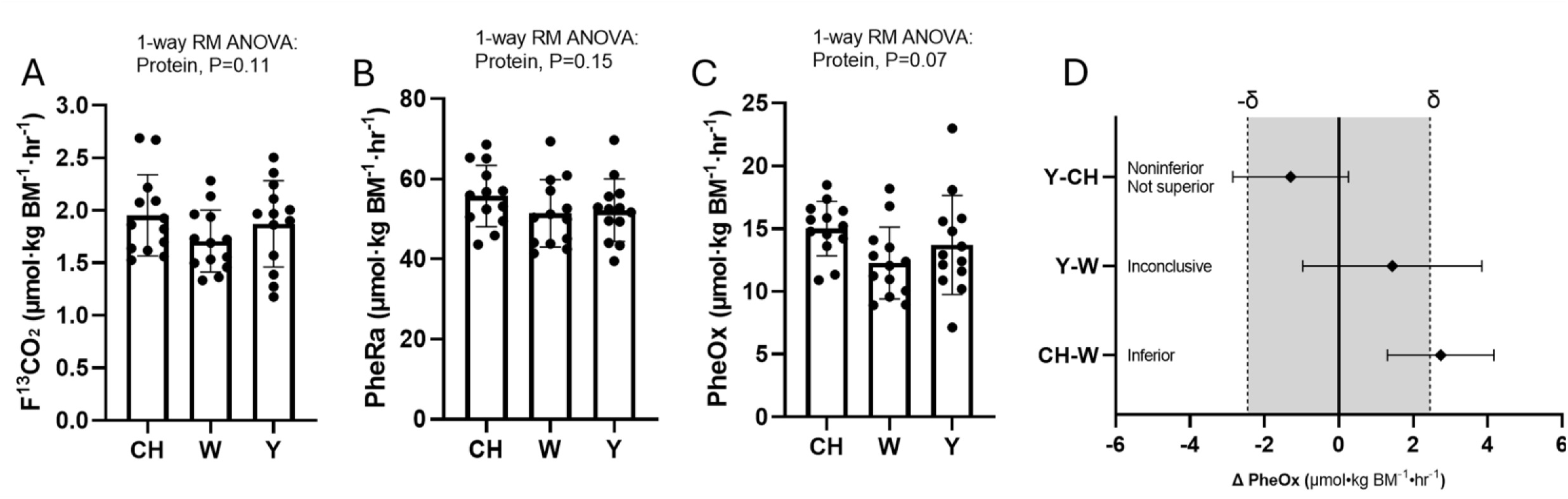
Complete phenylalanine kinetics and noninferiority data, inclusive of outlier trials. (**A**) Fraction of expired ^13^CO_2_ (μmol·kg BM^−1^·hr^−1^), (**B**) rate of phenylalanine appearance (PheRa) (μmol·kg BM^−1^·hr^−1^), (**C**) rate of phenylalanine oxidation (PheOx) (μmol·kg BM^−1^·hr^−1^), and (**D**) Noninferiority of protein sources (whey (W), collagen hydrolysate (CH), and yeast (Y)) given change in PheOx. Data are presented as means ± SD (n = 13 per group). (A-C) Data are analyzed by a 1-way repeated-measures ANOVA. There was no main effect of protein sources on F^13^CO_2_, PheRa, or PheOx, although protein source trended to a main effect on PheOx (P=0.07). (D) Mean differences are analyzed by one-tailed t-test, with W or CH set as the control. Noninferiority against W or CH is accepted when the upper bound of the 90% CI in change of PheOx is lower than the predetermined 20% noninferiority margin, with superiority accepted when lower bound crosses the superiority margin (+/-δ, +/-2.45 μmol·kg BM^−1^·hr^−1^, *grey shading*), with inconclusivity reached when the lower and upper bound spread across no difference in PheOx and the noninferiority margin.

## Notes

### Clinical Trial

NCT07148908

### Author Declarations

Ethics committee of the University of Toronto gave ethical approval for this work. All participants were informed of the purpose of the study, experimental procedures, and potential risks and gave written informed consent. The studies were performed in accordance with the Declaration of Helsinki, and the study protocols were approved by the University of Toronto Delegated Research Ethics Board (REB#00048239). The study is registered at clinicaltrials.gov (NCT07148908).

